# Evaluation of epigenetic and metabolomic biomarkers indicating biological age

**DOI:** 10.1101/2022.12.05.22282968

**Authors:** Lieke M. Kuiper, Harmke A. Polinder-Bos, Daniele Bizzarri, Dina Vojinovic, Costanza L. Vallerga, Marian Beekman, Martijn E.T. Dollé, Mohsen Ghanbari, Trudy Voortman, Marcel J.T. Reinders, W.M. Monique Verschuren, P. Eline Slagboom, Erik B. van den Akker, Joyce B.J. van Meurs

## Abstract

Biological age captures a person’s age-related risk of unfavorable outcomes using biophysiological information. Multivariate biological age measures include frailty scores and molecular biomarkers. These measures are often studied in isolation, but here we present a large-scale study comparing them.

In two prospective cohorts (*n*=3,196), we compared epigenetic (DNAm Horvath, DNAm Hannum, DNAm PhenoAge, DNAm GrimAge) and metabolomic-based (MetaboAge, MetaboHealth) biomarkers in reflection of biological age, as represented by five frailty measures and overall mortality.

We observed that mortality-trained biological age markers, DNAm GrimAge and MetaboHealth, outperformed age-trained biomarkers in frailty reflection and mortality prediction. The associations of DNAm GrimAge and MetaboHealth with frailty and mortality were independent of each other and of the frailty score mimicking clinical geriatric assessment.

Epigenetic, metabolomic, and clinical biological age markers seem to capture different aspects of aging. These findings suggest that mortality-trained molecular markers may provide novel phenotype reflecting biological age and strengthen current clinical geriatric health and well-being assessment.

---

Age is the most prominent risk indicator for common chronic diseases, frailty, and mortality(1–3). However, not everyone ages at the same rate. There are large interindividual differences in the biological aging process and rate of functional decline. Hence, the field of ageing research is in need of standardized markers that reflect the biological age of individuals and can provide phenotypes reflecting the ageing rate to be studied in depth. Besides, geriatric specialists use the comprehensive geriatric assessment (CGA) to identify the medical, social, and functional needs of older patients(4). This generic health assessment is, for instance, used in the clinic to determine whether elderly patients are fit enough to undergo invasive treatments(5) and is considered the gold standard for the treatment of frail patients(4). The CGA is time- and resource-consuming and mainly narrative-based(4). So far, there is no consensus for a multivariate molecular biomarker to capture the entire complexity of the aging process faithfully which could be used as a phenotype of biological age in ageing research and provide an indicator of overall health in the clinic.

Consensus is lacking on the operationalization of frailty in research practice, leading to the introduction of a variety of frailty measures(2,6–10). The Multidimensional Prognostic Index (MPI) is directly derived from the CGA and is regarded as the most direct translation of the CGA for research purposes. The MPI contains information on the medical, social, and functional status of the participants(9). The frailty phenotype (FP), also known as Fried frailty(2), is the most commonly used frailty score to assess physical frailty and has recently been translated into a continuous score, Continuous Fried Frailty (Cont.)(7). The Tilburg Frailty Indicator (TFI) combines the physical domain with the psychological and social domain(8). The frailty index (FI) measures frailty as an accumulation of deficits over a wide range of health domains(11).

Furthermore, several attempts have been made to capture the discrepancy between an individual’s chronological age and their age based on biological and clinical information, known as biological age, in a biomarker. In the past two decades, large-scale molecular data were used to develop several molecular markers of biological age based on, for example, telomeres, DNA methylation (DNAm), and metabolomics(12). Well-known are the DNAm or epigenetic aging clocks. These epigenetic aging clocks are biomarkers based on methylation values at a combination of specific CpG sites by which chronological age is best reflected. The first-generation epigenetic aging clocks, DNAm Horvath(13) and DNAm Hannum(14), were trained on chronological age and outperformed other aging biomarkers, such as telomere length, in the reflection and prediction of the aging process(12). Since physiological deficits resulting from and contributing to aging do not develop in a regular, clock-like manner, the second-generation epigenetic aging biomarkers were developed by training CpG-models based on a multi-system proxy of physiological dysregulation (DNAm PhenoAge(15)) or mortality risk (DNAm GrimAge(16)).More recently, metabolomics-based aging biomarkers were established using a well-standardized nuclear magnetic resonance platform(17,18). These biomarkers of biological age were trained on either chronological age (MetaboAge(19)) or mortality (MetaboHealth(20)).

Previous studies have shown that DNAm PhenoAge and DNAm GrimAge outperform the first-generation epigenetic aging biomarkers in reflecting physical health outcomes, cognitive and physical capacity, and prediction of overall mortality(21,22). However, the performances of the newly developed metabolomic biological age biomarkers have not been compared with either the first or second-generation epigenetic aging biomarkers. Moreover, whether epigenetic and metabolomic aging biomarkers capture different aspects of the aging process is unknown. Lastly, it is unclear whether aging biomarkers have added value to the CGA and, thus, their possible clinical applicability.

The current study compares the reflection of biological age of the first and second-generation epigenetic and metabolomic aging biomarkers by determining their association with five different frailty scores and with mortality. These outcomes largely reflect the aging process.

## Methods

### Study cohorts

The current study is a nested cohort study of data from the second and third cohort of the population-based Rotterdam Study (RS) and the second generation of the Leiden Longevity Study (LLS). In the Rotterdam Study, all residents of Ommoord, a suburb of Rotterdam, above the age of 55 years were invited to participate (23). Inclusion criteria were having complete information on all epigenetic biomarkers of biological aging based on either 450K-data (n=687) or EPIC-data (n=737), metabolomics biomarkers of biological aging, cell counts, and body mass index (BMI), leaving the study population to 1,424 participants. Ten of these participants withdrew their consent for longitudinal follow-up but wanted to participate in cross-sectional studies. We used their information for the frailty analyses but not for the mortality analyses.

The LLS cohort consists of 420 Dutch Caucasian families with at least two long-lived full siblings who were alive and willing to participate in the study.(24) Complete data on metabolomics, BMI, and mortality information was present in 1,849 LLS participants of the children of the long-living family members. In a subcohort of 584 participants, information on DNAm was available. Together these participants represent the study population for the external validation of our findings. A more detailed description of the cohorts can be found in the Online-Only Methods.

### DNA methylation

Genome-wide DNA methylation data was obtained from whole blood. In 687 RS participants and the LLS, we analyzed the samples using Illumina Infinium Human Methylation 450 K (450K) array(24,25). In the other 737 RS samples, we used Illumina Infinium MethylationEPIC BeadChip v1 manifest B5 (EPIC) arrays(26). The quality control procedures are described in the Online-Only Methods.

### Metabolomics

Metabolomic biomarkers from EDTA plasma were measured using high□throughput NMR metabolomics (Nightingale Health Ltd., Helsinki, Finland; biomarker quantification version 2016)(18). This technique quantifies over 200 metabolic measures, including routine lipids, lipoprotein subclass profiling with lipid concentrations within 14 subclasses, fatty acid composition, and various low-molecular-weight metabolites in molar concentration units(17,18).

### Biomarkers of biological aging

We calculated DNAm Horvath(13), DNAm Hannum(14), DNAm PhenoAge(15), and GrimAge(16) using R and Python scripts provided by the researchers who developed these measures and the R methylclock package(27). In case of the 3,332 CpG sites were information needed to calculate the biological ages was absent on the EPIC array, we imputed them with their mean value in the GOLD consortium, as described previously(28). The metabolomic biomarkers were used to compose MetaboAge(19) and MetaboHealth(20). MetaboAge was calculated using MiMIR(29), the dedicated R-shiny package, on the raw metabolomic biomarkers(19). To calculate the MetaboHealth, we log-transformed the fourteen biomarkers of interest and multiplied them by the natural logarithm of their hazard ratio on mortality in the fully adjusted joint model, as reported in the original manuscript(20). Finally, we calculated the chronological age independent part of the above-mentioned variables that we defined as the biomarkers of biological aging to use in all analyses in this study. We did so by taking the residual from the linear regression model of chronological age on the before-mentioned epigenetic and metabolomic variables. As we use the age-independent part, all biological aging biomarkers have, per definition, no correlation with chronological age.

### Assessment of mortality

Based on a linkage with the mortality registry of the municipality and the digitally connected medical records of the GPs working in the study area, we gathered information on the vital status of the participants on a bimonthly basis(30). The information on the vital status of participants in Rotterdam was last updated on the 20^th^ of October 2022.

The vital status of the participants of the LLS was updated in January 2021 through the Personal Records Database, which is managed by the Dutch governmental service for identity information(31).

### Frailty assessment

We used interviews, physical examinations, blood sampling, and general practitioners’ records to obtain information on the participants’ frailty. Using this information, we constructed the frailty phenotype (FP)(2), continuous Fried (Cont.)(7), the Frailty Index (FI)(6), the Tilburg Frailty Indicator (TFI)(32), and the Multidimensional Prognostic Index (MPI)(9). A more detailed description of the construction of these five frailty measures and the in literature described cut-offs to classify participants as either frail or non-frail can be found in the Online-Only Methods.

### Assessment of covariates

A questionnaire at baseline provided information on the sex and chronological age at blood sampling for all participants. We weighted and measured participants when they visited the research center; based on this information, BMI was calculated (kg/m^2^). We classified participants as smokers or non-smokers based on the answer to the question: “are you currently smoking?”. We defined cell counts as the measured white blood count percentage of lymphocytes and monocytes, making the percentage of granulocytes a given.

### Statistical analysis

The biomarkers of biological aging were, as before mentioned, defined as the residual of a linear regression of chronological age on the epigenetic and metabolomics measures. Therefore, these biological aging biomarkers were constructed per dataset. We determined the correlation between the biomarkers of biological aging and the correlation between the frailty scores using Spearman’s log rank correlation. To increase the homoscedasticity, one of the assumptions of linear regression analyses, we transformed the frailty indices with a Yeo-Johnson transformation using the *bestNormalize* R-package(33). We decided upon using Yeo-Johnson transformations as it allows using zeroes in the transformation(33) since having a frailty score of zero was valuable information that we did not want to lose in the power transformation. Linear regression models were used to determine the association between cross-sectional continuous Yeo-Johnson transformed Z-scored frailty Z-scored and biological aging biomarkers. Standardization was performed in the subcohorts separately for the subcohort analyses and in the combined information on all participants for the analyses involving the overall study population. We used Z-scores of the continuous outcomes as well as the aging predictors to be able to compare the effect sizes. We determined the association between a biomarker of biological age and cross-sectional binary frailty outcomes with logistic regression analyses. The linear and logistic regression analyses were adjusted for chronological age at blood sampling, sex, cell counts, BMI, and visit and cohort within the Rotterdam Study. The analyses in the entire RS study population were additionally adjusted for underlying subcohort and, thereby, methylation array and metabolomics batch used. Furthermore, we performed a sensitivity analysis in which we additionally adjusted for smoking status.

We used the R-package *survival* to compose Cox Proportional hazards regression models with chronological age at blood sampling to determine the association of the standardized aging predictors with overall mortality. We performed the analyses in four models in the RS and validated only the first two models in LLS as the MPI was not available in the LLS. The first model adjusted for sex, cell counts, BMI, and study-specific covariates. In the second model, we additionally adjusted for the smoking status. In the third model, we adjusted for the same covariates as in the first model, but we additionally adjusted for the MPI. The MPI mimics the CGA, as used in the clinic, best. Adjusting for the MPI provides an opportunity to determine the value of aging predictors beyond ongoing practice. Fourthly, we did a sensitivity analysis in which we additionally adjusted the third model for smoking status. Furthermore, we determined the association between frailty measures per standard deviation increase and overall mortality with a Cox Proportional Hazard model with time contributed to the study as the timescale. We did not use age as the timescale since some frailty measures, such as the frailty index, were, per definition, correlated with chronological age (Appendices 1-2). We adjusted the analyses for chronological age, BMI, cell counts, and sex. We again performed a sensitivity analysis in which we additionally adjusted for smoking status.

We performed all analyses in R version 4.1.3. Fig. 1 was created with BioRender.com.

**Figure 1.**
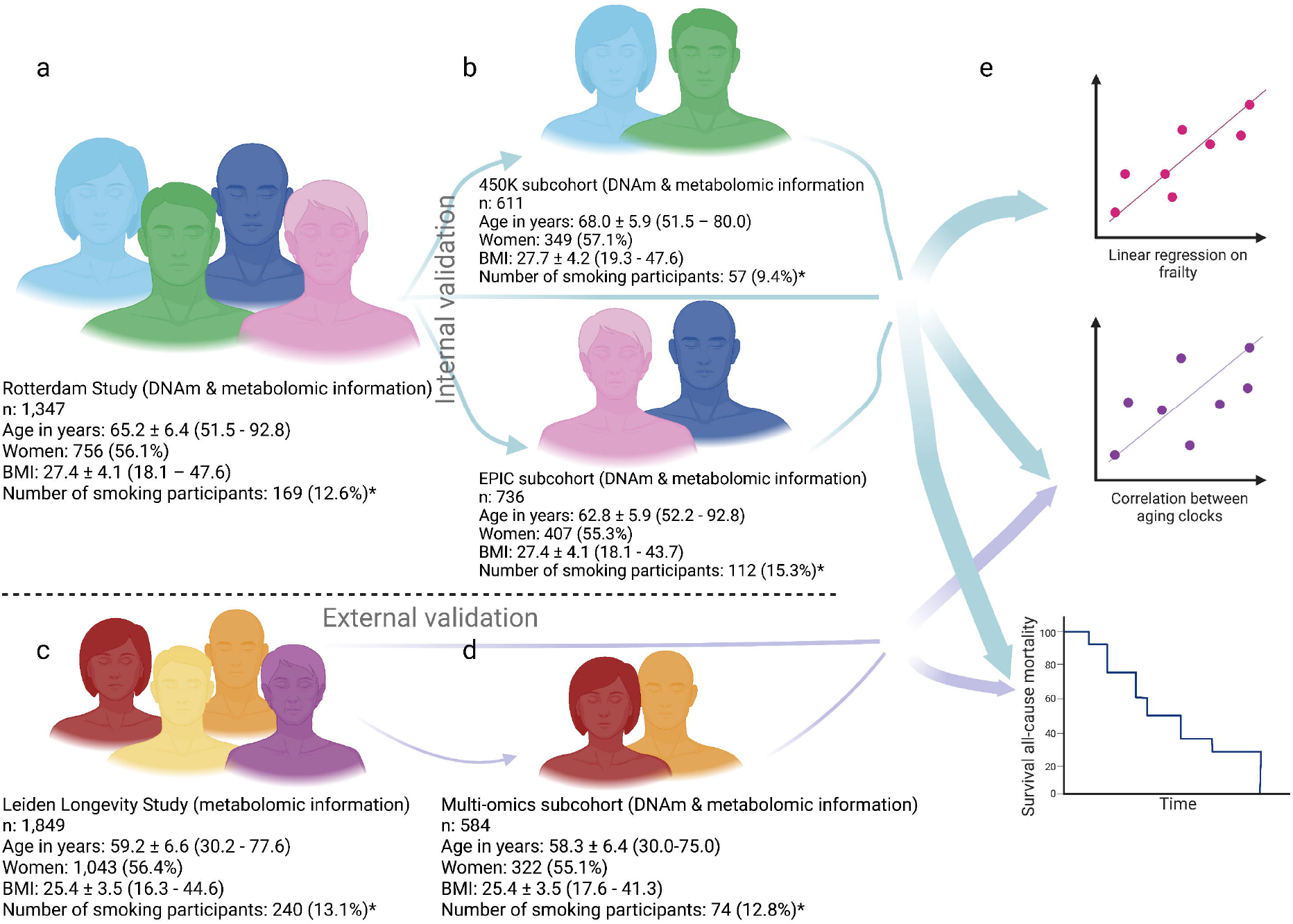
Outline of the study and study population characteristics * Smoking status was unknown for 5 Rotterdam Study participants (3 participants of the 450K-subgroup, 2 participants of the EPIC-subgroup) and for 17 LLS participants of whom 4 belonged to the multi-omics subgroup **a**. The Rotterdam Study overall study population with population characteristics; **b**. the Rotterdam Study two subcohorts, 450K and EPIC, stratified by the DNA methylation array used and their population characteristics; **c**. The external validation cohort, the Leiden Longevity Study with population characteristics; **d**. The subcohort of the Leiden Longevity Study where epigenetic information was available with population characteristics; **e**. We used both the overall Rotterdam Study population as its two subcohorts 1) to determine the correlations between each of the biological aging biomarkers, 2) to perform a linear regression for the association between the biological aging biomarkers and frailty, and 3) to determine the association between each of the biological aging biomarkers and all-cause mortality. We externally validated the correlations between the biological aging biomarkers and the association between the biological aging biomarkers and all-cause mortality in the Leiden Longevity Study and its subcohort. In **a-d**, BMI indicates body mass index; DNAm, DNA methylation; and n, size of the study population. Population characteristics in **a-d** are shown as a number for the population size; mean ± standard deviation (range) for age and BMI; and number (percentage) for the number of women and the number of participants currently smoking.

## Results

We used two distinct cohorts for our analyses: the Rotterdam Study (RS)(23), a population-based study, and the Leiden Longevity Study (LLS)(34), a long-living family study (Figure 1, eTable 1). The RS was separated into two sub-cohorts where the distinguishing factor was the DNAm array used, either 450K (*n=*611) or EPIC (*n=*736). For the LLS, the study population consisted of all participants with metabolomics information (n=1,849) with a multi-omics subcohort (n=584) of offspring and their partners from families without a family history of longevity, thereby selecting a subcohort closest to the population at large(35). We calculated the biomarkers of biological aging as the age-independent part of the aging biomarkers and used this metric in all further analyses (Methods: Biomarkers of biological aging).

### Correlation between biological aging biomarkers

Spearman’s rank correlation coefficients between the four epigenetic and two metabolomic aging biomarkers were low to moderate, ranging between 0.04 and 0.50 (Figure 2a). The highest correlation was found between DNAm Hannum and DNAm PhenoAge. The metabolomic aging biomarkers had the highest correlations with each other and with DNAm GrimAge. The correlations between biomarkers trained on chronological age from different molecular origins, i.e., DNAm Horvath or DNAm Hannum versus MetaboAge, were low, namely 0.04 between MetaboAge and DNAm Hannum and 0.16 between MetaboAge and DNAm Horvath. Interestingly, these correlations were lower than the observed correlation of 0.20 between MetaboAge and the mortality-trained DNAm GrimAge. The mortality-trained aging biomarkers DNAm GrimAge and MetaboHealth, trained on respectively DNAm and metabolomics, had a correlation of 0.30. Correlation patterns were similar across different methylation arrays (within RS) and across the two cohorts (RS and LLS) (eFigure 1)

**Figure 2.**
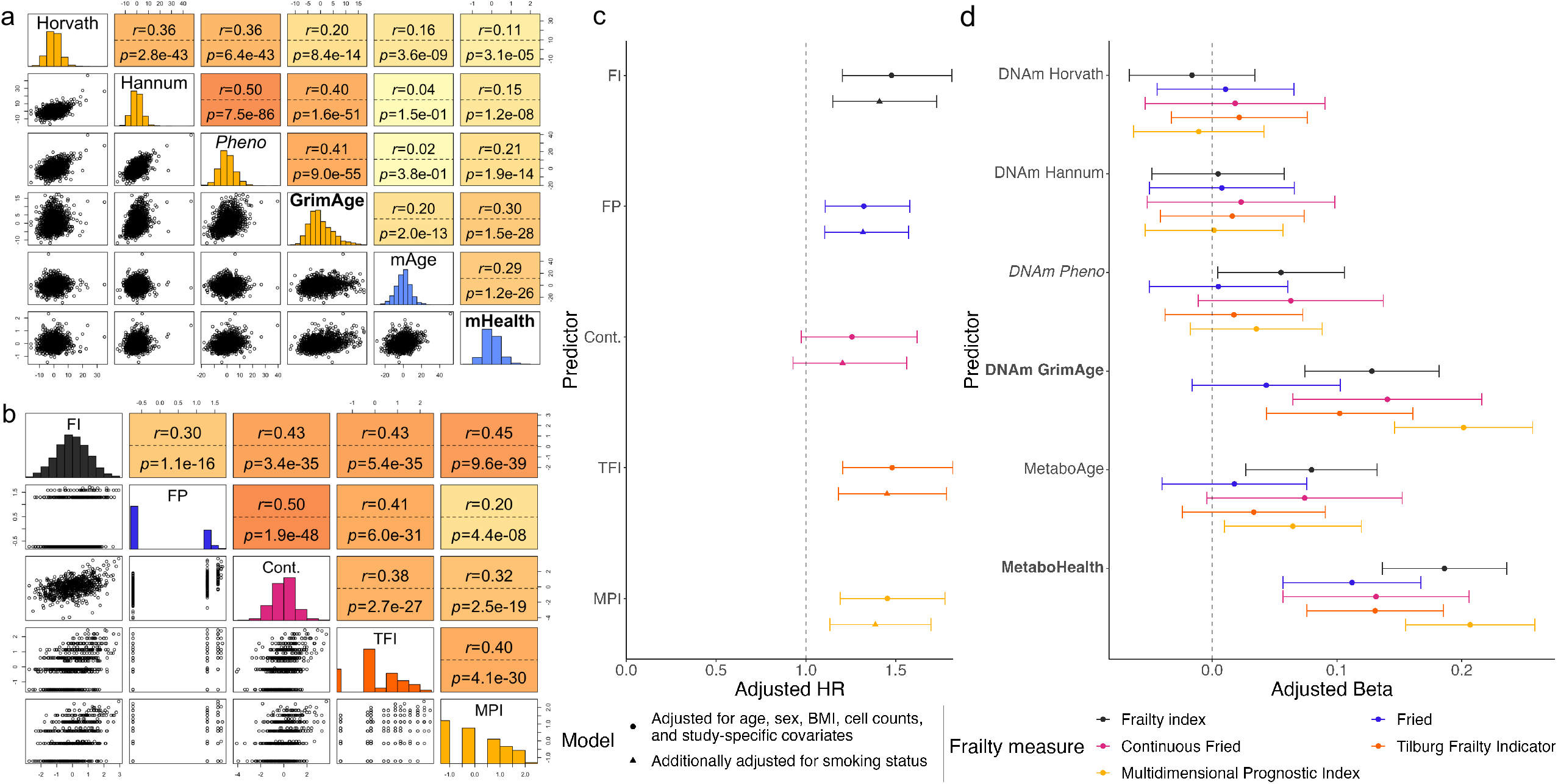
Correlations between biological age measures and the association between biomarkers of biological aging and frailty **a**. Spearman’s correlation of the different biological aging biomarkers in 1,424 Rotterdam Study participants with the histograms of epigenetic aging biomarkers in yellow and metabolomic-based aging biomarkers in blue. Labels in bold indicate a mortality-trained aging biomarker; cursive label a phenotypic age-trained aging biomarker; and the regular font an aging biomarker trained on chronological age. Values after *r=* represent Spearman’s rank coefficient; values after *p=* represent the p-value; the background color is darker for higher correlations. Horvath indicates DNAm Horvath; Hannum, DNAm Hannum; Pheno, DNAm PhenoAge; GrimAge, DNAm GrimAge; mAge, MetaboAge; and mHealth, MetaboHealth; **b**. Spearman’s correlation between the different Yeo-Johnson-transformed frailty measures in the 746 Rotterdam Study participants with information on all five frailty measures.. Values after *r=* represent Spearman’s rank coefficient; values after *p=* represent the p-value; the background color is darker for higher correlations. FI indicates frailty index, FP: frailty phenotype, Cont.: continuous Fried, TFI: Tilburg Frailty Indicator, and MPI: Multidimensional Prognostic Index; **c**. Risk of all-cause mortality per standard deviation increase of the Yeo-Johnson transformed FI(*ncases=*130/*n=*1330), FP(*ncases=*132/*n=*1328), Cont.(*ncases=*69/*n=*743), TFI(ncases=129/*n=*1328), MPI(*ncases=*132/*n=*1333) in the RS overall study population. The figure represents the adjusted hazard ratios and 95%-confidence intervals. **d**. Associations of standardized biological aging biomarkers with standardized FI(*n=*1,341), FP(*n=*1,339), Cont.(*n=*748), TFI(*n=*1,339), and MPI(*n=*1,344) based on linear regression analyses in all participants of whom data on biological aging biomarkers and frailty were available in the overall Rotterdam Study dataset. Analyses were adjusted for age, sex, BMI, cell counts, subcohort, and Rotterdam Study cohort and visit. The figure represents the adjusted betas and 95%-confidence intervals.

### Frailty

Frailty measures, like the biological aging biomarkers, were developed to capture the individual aging process(11,12,36); they represent measures of biological age. To determine whether different frailty measures developed based on different rationales could be used interchangeably, we measured five different frailty scores in our study population: the FI(6), the FP(2), Cont.(7), the TFI (8,32), and the MPI(9) (Online-Only Text 1 & 2). When elements from these frailty measures were lacking in our dataset, we used proxies (Online-Only Text 2).

As shown in Figure 2b, the correlation between the different frailty measures ranged from 0.22 to 0.52. The highest correlation was observed between the two physical frailty measures FP and Cont. and the lowest between FP and the MPI, the latter being the frailty measure directly derived from the CGA. All frailty measures were associated with an increased risk on overall mortality. We observed higher hazard ratios for frailty scores including information beyond the physical domain than the physical frailty measures, yet this difference was not statistically significant (Figure 2c, eTable 2).

We concluded that the frailty measures could not be used interchangeably. Therefore, we examined the six biological aging biomarkers (four epigenetic and two metabolomic aging biomarkers) for their association with all five frailty measures. All biological aging biomarkers and the frailty scores were standardized to improve comparability. We observed that an increase in the biological aging biomarkers originally trained on mortality (DNAm GrimAge and MetaboHealth) was consistently associated with higher frailty scores in linear regression analyses adjusted for age, sex, body mass index, cell counts, and study-specific covariates. Moreover, these results were more prominent than we observed for biological aging biomarkers trained on chronological age (clocks), with MetaboAge among the clocks showing the most prominent association with frailty scores, and phenotypic age. The strongest associations were found for MetaboHealth (adjusted beta in the RS combined study population per standard deviation increase (B 0.20 95%-confidence interval [CI] [0.15;0.25]) and DNAm GrimAge (B 0.21 [CI 0.16;0.26]) with the MPI (Figure 2d, eFigure 2, eTable 3). The analyses were adjusted for age despite using chronological age-independent aging biomarkers to address the inherent correlation of frailty measures, for example the frailty index, with chronological age. BMI was included as a covariate as both epigenetic-(37) and metabolomic-based(19) aging biomarkers are known to associate with a higher BMI and BMI information is included in all frailty scores(2,6–9) (Online-Only Text 1 & 2). We performed a sensitivity analysis adjusting for smoking to determine whether the inclusion of smoking-pack years in the construction of the DNAm GrimAge was driving the results. The sensitivity analysis did not remarkably alter the results (eTable 3).

Subsequentially, we determined whether the associations with the frailty measures of the best-performing epigenetic and best-performing metabolomic aging biomarker were independent of each other. Both DNAm GrimAge and MetaboHealth remained independently associated with frailty in a linear regression adjusted for the same covariates as the univariable analyses (Table 1, Extended Data 6: Extended Table 4). There were some small improvements of the explained variance of the models when both DNAm GrimAge and MetaboHealth were included, for example the explained variance of the association with frailty index improved from 0.22 for DNAm GrimAge and 0.23 for MetaboHealth to 0.24 in the combined model (eTable 3, eTable 4). Additionally adjusting for smoking status did, again, not considerably change the results (eTable 4). Furthermore, the same pattern appeared when categorizing participants as non-frail and frail using the traditional cut-offs (Online-Only Text 1) of the frailty measures (eTable 5).

**Table 1.** Results of multivariable regression models including both DNAm GrimAge and MetaboHealth as exposures and frailty measures as outcome

|  |  | DNAm GrimAge |  | MetaboHealth |  |
| --- | --- | --- | --- | --- | --- |
|  |  | Adjusted* Beta | P-value | Adjusted* Beta | P-value |
|  |  | per SD (CI) |  | per SD (CI) |  |
| FI | n=1,341 | 0.08 (0.03;0.14) | 4.27x10<sup>-3</sup> | 0.17 (0.11;0.22) | 4.02x10<sup>-10</sup> |
| FP | n=1,339 | 0.01 (-0.05;0.07) | 0.69 | 0.11 (0.05;0.17) | 2.02x10<sup>-4</sup> |
| Cont. | n=748 | 0.11 (0.03;0.19) | 5.39x10<sup>-3</sup> | 0.10 (0.02;0.18) | 0.01 |
| TFI | n=1,339 | 0.07 (0.01;0.13) | 0.02 | 0.11 (0.06;0.17) | 9.88x10<sup>-5</sup> |
| MPI | n=1,344 | 0.15 (0.10;0.21) | 1.20x10<sup>-7</sup> | 0.17 (0.12;0.22) | 6.67x10<sup>-10</sup> |

\* Adjusted for chronological age at blood sampling, sex, body mass index, cell counts, subcohort, and visit and cohort within the Rotterdam Study.
SD indicates standard deviation; CI, 95%-confidence interval; FI, frailty index; FP, frailty phenotype; Cont., continuous Fried; TFI, Tilburg Frailty Indicator; MPI, multidimensional prognostic index; and n, number of participants for whom we have information on this frailty score.

### Mortality

Beyond reflecting on an individual’s current state, biomarkers of biological age are believed to capture predictive information on the aging process(12). We, therefore, determined the association of biological aging biomarkers with mortality during 11,281 person-years of follow-up in the RS. The median follow-up time was 8.6 years. During follow-up, 132 participants died. A higher DNAm GrimAge and MetaboHealth were associated with a higher risk of overall mortality in both subgroups as well as the combined study population. The highest risk estimates for all-cause mortality were observed for DNAm GrimAge (adjusted hazard ratio in the RS combined study population per standard deviation increase (HR) 1.79 95%-confidence interval [CI] [1.52;2.12]) and MetaboHealth (HR 1.79 [CI 1.52;2.09]) (Figure 3a, eFigure 3, eTable 6). Yet, the observed hazard ratios were a bit more stable for DNAm GrimAge than for MetaboHealth. The Cox Proportional Hazard models were adjusted for the same covariates as used in the linear regression except for age, which was included in the timescale. A sensitivity analyses to determine whether smoking status influenced the risk entailed by the aging biomarkers, again, did not noteworthy shift the results.

**Figure 3.**
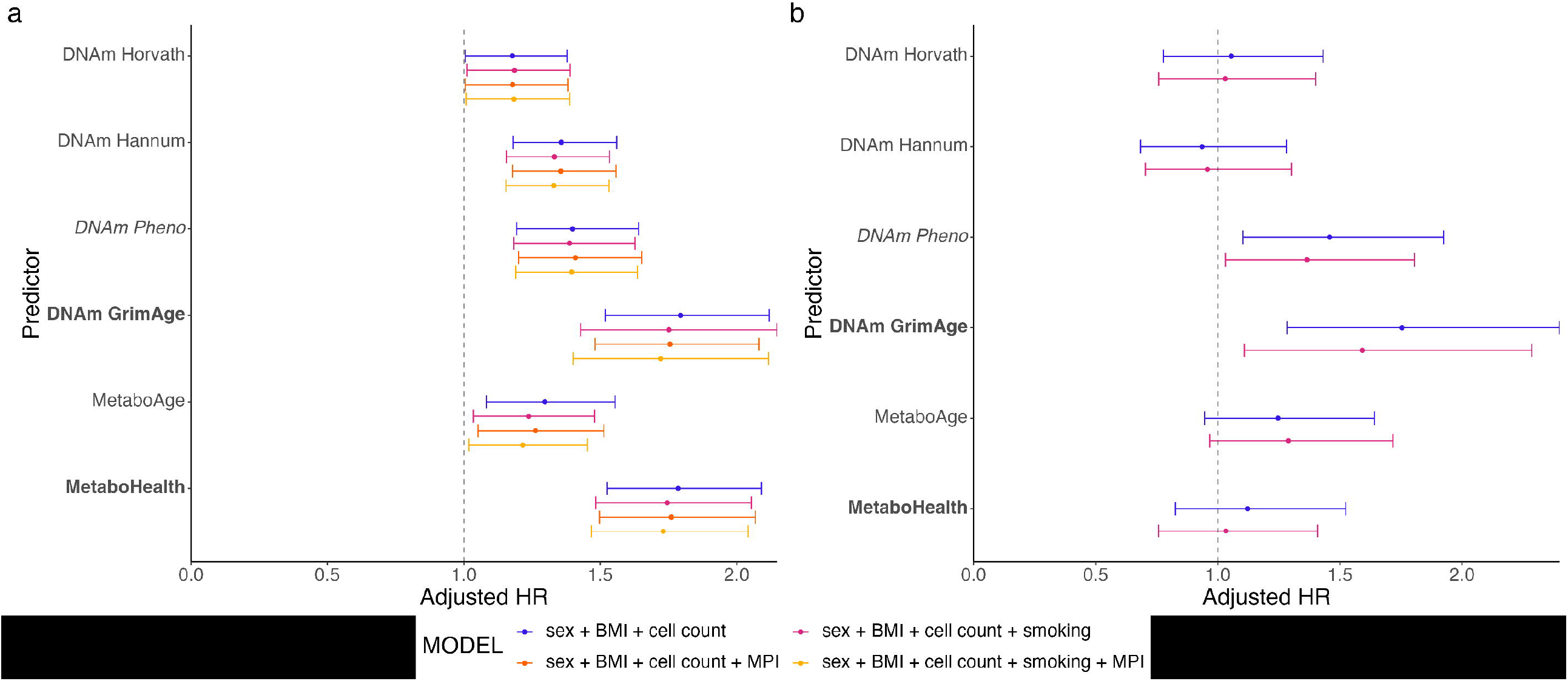
Aging predictors and their univariable risk of all-cause mortality per SD Risk of all-cause mortality per standard deviation increase of the aging predictors in **a**. the overall Rotterdam Study population (n=1,336); and **b**. the subgroup of the Leiden Longevity Study with information on the epigenetic aging predictors (n=584). BMI indicates body mass index; HR, hazard ratio; and MPI, multidimensional prognostic index.

We then assessed whether the observed associations between the aging biomarkers and mortality were explained by frailty. For this, we used the MPI since it is the frailty score most closely related to the CGA. Earlier we showed that the MPI itself is associated with an increased risk of overall mortality (Figure 2c, eTable 2). Nevertheless, the associations between the aging predictors and mortality were independent of and not notably changed by the MPI (Figure 3a, eFigure 3, eTable 6). These results remained, yet again, unchanged in a sensitivity analysis adjusting for smoking status (Figure 3a, eFigure 3, eTable 6).

To determine whether the risk of mortality captured by the best performing epigenetic aging predictor and metabolomic aging predictor were mutually independent, we performed a Cox-proportional hazard analysis including both aging predictors and adjusted for sex, BMI, and cell count. When combining DNAm GrimAge and MetaboHealth in a model, both showed an independent risk of all-cause mortality, respectively DNAm GrimAge (HR 1.56 [CI 1.31;1.85]) and MetaboHealth (HR 1.60 [CI 1.35;1.89]). The concordance increased slightly from 0.69 when only using DNAm GrimAge and 0.67 when only including MetaboHealth to 0.70 in the combined model. These results were robust among the RS overall study population and subcohorts. These results remained, again, similar when adjusting for the MPI and smoking status (Table 2, eTable 7).

**Table 2.**
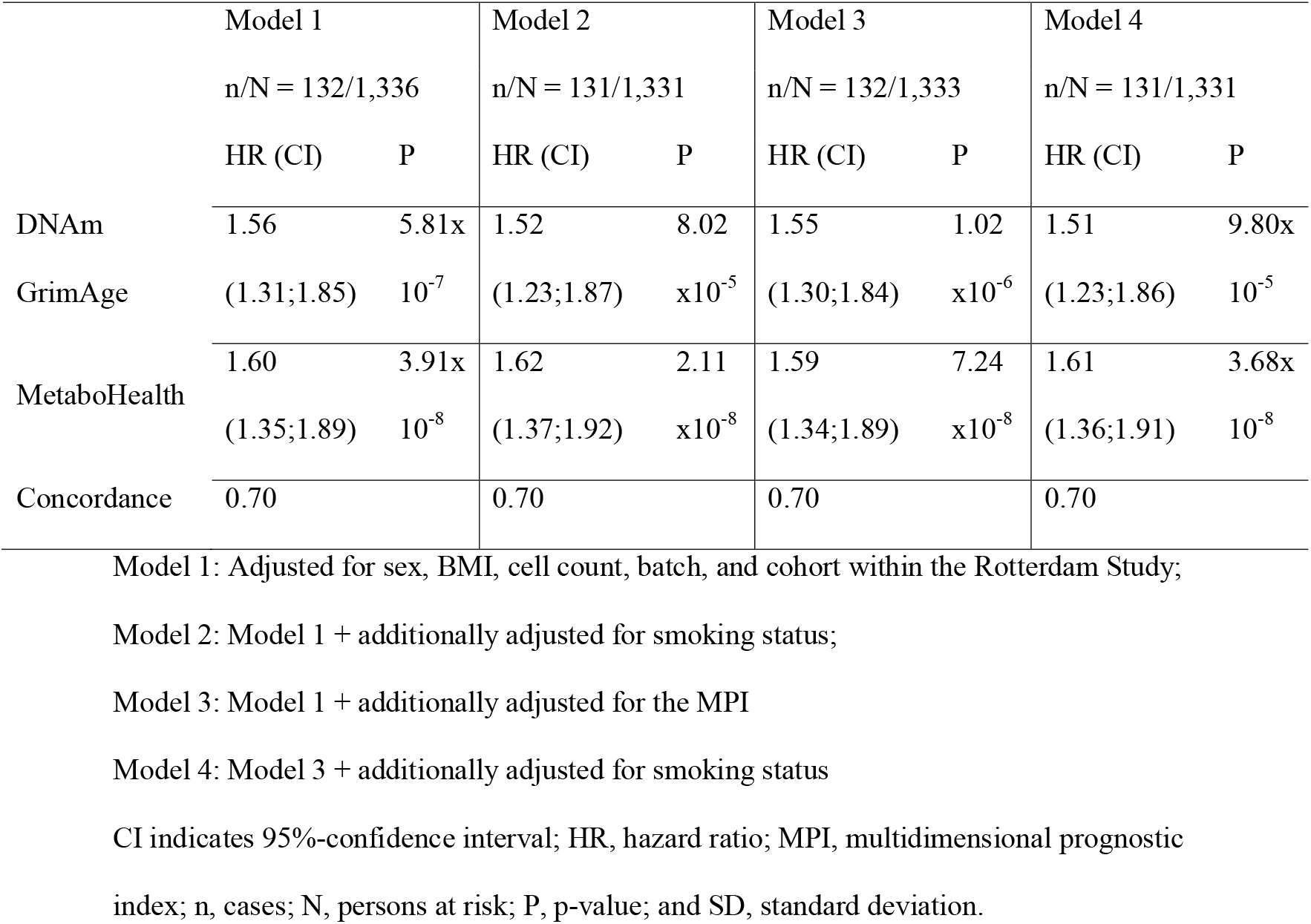
The multivariable risk of all-cause mortality of DNAm GrimAge and MetaboHealth

In our external validation cohort, the LLS, 147 participants died during follow-up (median 13.3 years), of whom 43 participants were part of the multi-omics subcohort that represented 7,553 (median 13.2 years) of person-year follow-up. In the subcohort, where we could validate both the epigenetic and the metabolomic aging predictors, the results of DNAm PhenoAge, DNAm GrimAge, and MetaboAge were similar to the results in the RS. By contrast, the associations between the other aging predictors, especially MetaboHealth, and mortality drops significantly (Fig. 3b, eTable 6). In the overall population of the LLS, MetaboHealth outperformed MetaboAge in the prediction of mortality, yet the mortality risk of both aging predictors was smaller than in the RS overall population and more comparable to the EPIC-subgroup (eFigure 3, eTable 6).

## Discussion

There is no consensus on which biomarkers that can be measured in human studies in standardized fashion reflect biological age. In the present study we considered frailty measures and mortality as phenotypes representing biological age. We compared five frailty measures in a population-based cohort and four epigenetic and two metabolomic biomarkers of biological age in two distinct cohorts of which the majority of older participants were not clinically frail. Our most prominent findings were: 1) the rather weak to moderate correlations between DNAm and metabolomics biological aging biomarkers especially between biomarkers from different origins trained on chronological age; 2) the outperformance of the mortality-trained biomarkers, MetaboHealth and DNAm GrimAge, in reflection of biological age, as represented by frailty and mortality, compared to age-trained biological age biomarkers and frailty measures; 3) the mutually independent associations between the mortality-trained biomarkers of biological age, DNAm GrimAge and MetaboHealth with frailty and mortality; and 4) the independence of the mortality association of the biomarkers of biological age from the MPI, the frailty measure directly derived from the CGA, and smoking. These findings stress that the different molecular markers of biological age complement each other in estimating frailty and mortality risk and potentially complement standardized health assessment in the clinical setting.

Similar to previous studies(9,38–43), we found an association between higher frailty scores and an increased risk of all-cause mortality for all frailty measures of interest. Unfortunately, despite the large variety of frailty measures, only a few of them have been directly compared in other studies. Most studies comparing different frailty measures focused on the frailty index and frailty phenotype(44–46). Our results align with previous studies reporting a higher risk on overall mortality for the frailty index than for the frailty phenotype(45,46). However, this difference was not significant in our study, which could be a result of lack of power. We are not aware of other studies comparing the risk estimates of frailty measures with the molecular biomarkers of biological age assessed in this study. Nonetheless, a previous study comparing the frailty index with a frailty index build using 40 different biomarkers showed that the biomarker-based frailty index was associated with a higher mortality risk than the frailty index in participants who are not clinically frail, like most of our participants. Furthermore, this study also emphasizes the added value of using biomarkers in frailty assessment as they report higher discriminative ability when both the biomarker-based frailty and frailty index are included in mortality prediction(44).

Previous studies focusing on the comparison between different epigenetic aging biomarkers have found comparable results. For example, the Irish Longitudinal Study on Ageing compared the four DNAm aging predictors used in the current study in a sample of 490 participants. Similar to the present study, results showed that DNAm GrimAge (the mortality-based biomarker) outperformed the other three epigenetic aging-based biomarkers in the reflection of physical health outcomes and prediction of all-cause mortality(21). Another study, including participants from three British cohorts, showed that associations between cognitive and physical capacity were present with DNAm GrimAge and DNAm PhenoAge, but absent when assessing DNAm Horvath or DNAm Hannum(22).

The performance of the metabolomic-based biomarkers of biological age trained on the basis of age acceleration and mortality has, to our knowledge, not been evaluated in other studies. In the two cohorts from the Rotterdam Study, the outperformance of the mortality-trained markers in reflecting frailty and predicting mortality was evident, as was the case for the comparison of metabolomics aging predictors in the LLS study. The lower performance of these markers in the LLS subgroup might be caused by the small sample size since in the original studies of this cohort(19,20) the metabolomic markers predicted adverse outcomes equally well in RS and LLS. Another possible explanation might be the difference in follow-up time between the two cohorts. Due to the small sample size, we cannot check the latter. In the RS, we observed somewhat unstable results for DNAm Horvath and MetaboAge, and to a lesser extent, MetaboHealth. This might be a result of the usage of the age-independent part of the aging biomarkers. We used the age-independent part as our biomarkers by regressing out chronological age. This approach is more vulnerable to outliers when fewer participants are included in the study. Further analysis of the performance of these biomarkers of biological age in small studies is recommended to determine their applicability in studies with smaller sample sizes. The molecular markers of biological age could potentially be used in the clinical setting to improve health and resilience estimates and as response monitors in intervention studies. In both future applications, the performance in limited sample-sized groups and ultimately even in individual patients is crucial.

Both MetaboHealth and DNAm GrimAge were trained on prospective mortality, MetaboHealth directly and DNAm GrimAge by creating DNAm surrogates of plasma proteins and smoking-pack years and using an elastic net Cox regression to the surrogates associated with overall mortality. DNAm PhenoAge was trained on phenotypic age, a predictor of mortality consisting of nine biomarkers and chronological age. Phenotypic age had a correlation with chronological age of 0.94 in NHANES IV(15). DNAm Horvath, DNAm Hannum, and MetaboAge were trained on chronological age. As MetaboHealth and DNAm GrimAge outclass the other aging biomarkers, not solely in mortality prediction but also frailty reflection, we believe training on mortality, or, for example, multimorbidity, improves the aging biomarkers’ ability to capture the heterogeneity in aging. Our findings align with the theory that fast-agers die sooner and consequently contribute less to the construction of biomarkers based on age, while biomarkers containing longitudinal information, such as mortality, suffer less from this selection bias(47). This finding could be of interest for future research aiming to create a new biological age biomarker. Based on the fact that DNAm GrimAge and MetaboHealth outperformed the other aging clocks in both RS subcohorts, as well as the LLS overall study population, we believe that our study provides further support for the benefits of training on longitudinal information regardless of the omics-layer used. This finding could have important implications for the development of future biomarkers as using longitudinal outcomes seems to results in aging predictors better equipped to capture the physiological heterogeneity that increases with aging. However, the performance of biological aging biomarkers on short-term outcomes still needs to be evaluated.

The low correlation of the epigenetic and metabolomic aging biomarkers in combination with the independent association of DNAm GrimAge and MetaboHealth with frailty and mortality suggests that metabolomic and epigenetic aging biomarkers capture different aspects of the aging process. However, there was only a slight increase in the explained variance when using both DNAm GrimAge and MetaboHealth. Furthermore, since the associations of these biomarkers of biological age were also independent of the MPI, there is an indication that these aspects are not captured in the CGA. This could imply that using these aging biomarkers would strengthen clinical geriatric risk assessment. Therefore, further research into the different aspects of aging captured by the different aging biomarkers and their applicability in the clinic would be advisable.

The main strengths of this study are the relatively large study population for which we had information on both DNAm and metabolomics, internal validation as well as external validation, and only a small loss to follow-up in the mortality data. Furthermore, with five different frailty measures, we had data on a wide variety of aspects of aging and were able to give insight into the distinct features of frailty measures. Besides, having information on both frailty and mortality gave us the opportunity to determine the associations of biological aging biomarkers with mortality adjusted for the MPI and, thereby, obtain an indication of the performance of molecular markers of biological aging beyond ongoing clinical practice.

However, there are some limitations of the current study. Firstly, participants needed to be fit enough to visit the research centers to provide blood samples and participate in several assessments for the frailty examinations. This requirement led to a selection bias towards healthy individuals. Secondly, the RS and the LLS were part of a large study in which the 14 biomarkers used in MetaboHealth were selected; of the 44,168 participants included in the construction of MetaboHealth, 13.7 percent originated from either LLS or RS. The contribution of both cohorts to the selection of MetaboHealth might have led to an overestimation of its association with frailty and mortality. Thirdly, we did not have all the original measurements on which the frailty measures are usually based. When a specific measurement was not present, we used proxies (Online-Only Text 2). We chose the proxies carefully with the help of a geriatrician; however, this could have had some impact on the estimations. Lastly, our study population consisted of white individuals aged 30 to 98 years; thus, validation of our project in other study populations and other (middle-aged) aspects of biological age is needed to assess the robustness of our results.

To our knowledge, this is the first study comparing the performance of both epigenetic and metabolomic-based aging biomarkers in reflecting frailty and mortality risk as measures of biological age. Furthermore, this is the first study including information on five different frailty measures as well as information on molecular biomarkers of biological age. We showed that epigenetic and metabolomic-based biomarkers of biological aging trained on mortality, DNAm GrimAge, and MetaboHealth reflected these biological age measures better than aging predictors trained on age or phenotypic age. The associations of DNAm GrimAge and MetaboHealth with frailty and mortality are independent of each other, suggesting that they capture information on different aspects of aging and may both be studied as novel phenotypes in research aimed at finding determinants of biological ageing. For the age and health category we have studied, it is also relevant that the associations of the biological age markers with mortality are partly independent of the MPI, a proxy for the standardized geriatric health assessment CGA as used in the clinic. These findings suggest that DNAm GrimAge and MetaboHealth could be valuable to complement the current health, well-being, and risk assessments in clinical practice. Therefore, further research into the potential integration of these biomarkers of biological aging in a clinical setting is warranted as well as increasing the informativity of these markers on the level of the individual patient.

## Supporting information

Online-Only

eTable 1

eTable 2

eTable 3

eTable 4

eTable 5

eTable 6

eTable 7

## Data Availability

Rotterdam Study data can be obtained upon request. Requests should be directed towards the management team of the Rotterdam Study, which has a protocol for approving data requests. For the data of the Leiden Longevity Study please contact Eline Slagboom or Marian Beekman or. Because of restrictions based on privacy regulations and informed consent of the participants, data cannot be made freely available in a public repository.

## Funding

The Rotterdam Study is supported by the Erasmus Medical Center and Erasmus University Rotterdam; the Netherlands Organization for the Health Research and Development (ZonMW); the Research Institute for Disease in the Elderly (RIDE); the Ministry of Education, Culture, and Science; the Ministry of Health, Welfare, and Sports; the European Commission; and the municipality of Rotterdam. The Leiden Longevity Study is supported by the European Union’s Seventh Framework Programme (FP7/2007-2011) under grant agreement number 259679. This study was financially supported by the Innovation-Oriented Research Program on Genomics (SenterNovem IGE05007), the Centre for Medical Systems Biology, and the Netherlands Consortium for Healthy Ageing (grant 050-060-810), all in the framework of the Netherlands Genomics Initiative, Netherlands Organization for Scientific Research (NWO), by BBMRI-NL, a Research Infrastructure financed by the Dutch government (NWO 184.021.007 and 184.033.111). The current study was supported by VOILA (ZonMW 457001001) and Medical Delta (scientific program METABODELTA: Metabolomics for clinical advances in the Medical Delta). EBvdA is funded by a personal grant from the Dutch Research Council (NWO; VENI: 09150161810095). METD is funded by the Ministry of Health, Welfare, and Sport & The National Institute for Public Health of the Netherlands (S/010003).

## Acknowledgements

The contribution of all participants of the Rotterdam Study and the Leiden Longevity Study is gratefully acknowledged; this research would not have been possible without them.

## Notes

### Competing Interest Statement

The authors have declared no competing interest.

### Author Declarations

Ethics committee/IRB of Leiden University Medical Center gave ethical approval for this work Ethics committee/IRB of Erasmus Medical Center gave ethical approval for this work

