## Supplementary material for "Evaluation of epigenetic and metabolomic biomarkers indicating biological age": Online-Only

Online-Only Materials

### Online-only Methods

#### Rotterdam Study

The current study is a nested cohort study of data from the second and third cohort of the population-based Rotterdam Study (1). In the Rotterdam Study, all residents of Ommoord, a suburb of Rotterdam, above the age of 55 years were invited to participate. The second cohort started in 2000 and included 3011 participants. In 2006, the third cohort of 3932 participants started. All participants had measurements at baseline and follow-up measures after 5-6 years. In the current study, we included people from the second cohort who came for their third re-evaluation to the research center and participants from the third cohort who visited the research center for the first or second time. Furthermore, inclusion criteria were having complete information on all epigenetic biomarkers of biological aging based on either 450K-data (n=687) or EPIC-data (n=737), metabolomics biomarkers of biological aging, cell counts, and body mass index (BMI), leaving the study population to 1424 participants. Ten of these participants withdrew their consent for longitudinal follow-up but wanted to participate in cross-sectional studies. We used their information for the frailty analyses but not for the mortality analyses.

The Medical Ethics Committee of the Erasmus MC (registration number MEC 02.1015) and by the Dutch Ministry of Health, Welfare and Sport (Population Screening Act WBO, license number 1071272-159521-PG) approved of the Rotterdam Study. The Rotterdam Study Personal Registration Data collection is filed with the Erasmus MC Data Protection Officer under registration number EMC1712001. All participants provided written informed consent to participate in the study and to obtain their information from treating physicians.

#### Leiden Longevity Study

The Leiden Longevity Study has been enrolled between 2002 and 2006. The LLS cohort consists of 420 Dutch Caucasian families with at least two long-lived full siblings who were alive and willing to participate in the study. Long-lived was defined as being at least 89 years old as a man or 91 years old as a woman. Besides the siblings (1st generation), the cousins (1st generation), the children (2nd generation), and the partners of the children (2nd generation) of the long-lived family members were recruited for the study. This resulted in the participation of 991 siblings and cousins (1st generation), 1,671 of their children (2nd generation), and 744 of the children’s partners (2nd generation). All participants had measurements at baseline and were followed up for mortality and morbidity(2). Complete data on metabolomics, BMI, and mortality information was present in 1,849 LLS participants of the 2nd generation. In a subcohort of 584 participants, information on DNAm was available. Together these participants represent the study population for the external validation of our findings.

In accordance with the Declaration of Helsinki, we obtained informed consent from all participants before they entered the study. Good clinical practice guidelines were maintained. The ethical committee approved the study protocol of the Leiden University Medical Center before the start of the study (P01.113).

#### DNA methylation

Genome-wide DNA methylation data was obtained from whole blood. In 687 RS participants and the LLS, we analyzed the samples using Illumina Infinium Human Methylation 450 K (450K) array(3,4). In the other 737 RS samples, we used Illumina Infinium MethylationEPIC BeadChip v1 manifest B5 (EPIC) arrays(5). In brief, all samples of DNA (500 ng of DNA per sample) were bisulfite converted using the Zymo EZ-96 DNA-methylation kit (Zymo Research, Irvine, CA, U69SA). The samples were hybridized according to the manufacturer’s protocols. The methylation proportion of a CpG site was reported as a β-value ranging between 0 (no methylation) and 1 (complete methylation). All methylation samples were processed at the Genetic Laboratory of Internal Medicine, Erasmus University Medical Centre, Rotterdam.

The quality control (QC) procedure for the 450K-subgroup and the LLS has been described in depth elsewhere(6,7). QC in the EPIC-subgroup was done using R version 4.0.5 and an R-pipeline consisting of the R-packages ewastools(8), minfi(9), and Meffil(10). We excluded 8 samples that failed technical controls (n=8), including extension, hybridization, and bisulfite conversion, according to the Illumina criteria and/or had a call rate <96% (n=2). Additionally, we checked for gender mismatches; this did not lead to additional exclusions. We excluded 45,079 unique probes with a detection p-value above the threshold of 0.01 or a bead number below 3 in at least 10% of the samples or that were known cross-reactive probes(5,11,12). The final number of probes after QC was 820,839. Then, we performed normalization of the QC passing probes beta values using the Quantile Normalization method based on six categories (QN-6C). We performed QN-6C using the CPACOR pipeline as explained in detail elsewhere(13). We applied this methodology as it has been suggested to outperform other approaches in the analysis of DNA methylation data where there is no expectation of global methylation changes among sample(13,14).

#### Frailty assessment

We used interviews, physical examinations, blood sampling, and general practitioners’ records to obtain information on the participants’ frailty. Interviews contained the Stanford Health Assessment Questionnaire to assess activities of daily living (ADL)(15), the Lawton Instrumental Activities of Daily Living scale to assess instrumental activities of daily living (IADL)(16), the Center for Epidemiologic Studies Depression Scale (CES-D) to assess depression(17), Mini-Mental State Examination (MMSE) to assess cognition(18)^,^ and questions on living situation, history of fractures, physical activity, socioeconomic status, smoking status, and medical history. Gait speed, anthropometric, blood pressure, and handgrip strength measurements were part of the physical examinations at the research center. At the research center, blood was collected to provide information on several plasma measurements, such as homocysteine, hormone levels, vitamin deficiencies, and cholesterol. We used general practitioner’s records to acquire information on prevalent and incident morbidity. Using this information, we constructed the frailty phenotype (FP)(19), continuous Fried (Cont.)(20), the Frailty Index (FI)(21), the Tilburg Frailty Indicator (TFI)(22), and the Multidimensional Prognostic Index (MPI)(23). A more detailed description of the construction of these five frailty measures and the in literature described cut-offs to classify participants as either frail or non-frail can be found in Online-Only Text 1.

### Online-Only Text 1

#### Frailty index

The frailty index (FI) conceptualizes frailty as the accumulation of health deficits(24). The construction of the FI in the Rotterdam Study has extensively been described elsewhere(21). In short, health-associated deficits with increased prevalence or severity with age that occurred in more than 5 percent and less than 80 percent of the participants were scored. When questions regarding deficits, for instance, from the same questionnaire, had a correlation above 0.7, only the question with the highest age correlation was used as a deficit. We scored each deficit a value between 0, absent, and 1, fully present. After multiple imputation, the sum deficit score was divided by the number of deficits. For the sake of robustness, we only included participants with information on at least 20 deficits.

#### Frailty phenotype

The frailty phenotype (FP) is a measure focusing on physical frailty(19,25). FP consists of five domains: weak grip strength, weight loss, exhaustion, slow walking speed, and low physical activity. A participant scored in each domain either a 0, not fulfilling this frailty criterium, or a 1, fulfilling this frailty criterium. We based all cut-offs on available literature(19,25).

Grip strength was measured using the highest value (kg) of three runs of a participant’s non-dominant hand on a handheld dynamometer. We defined weak grip strength in men as a grip strength lower than 29 kg with a BMI <24, or <30 kg given a BMI between 24.1 and 28, or <32 kg given a BMI >28, or in women <17.3 kg given a BMI <23, or <17.3 given a BMI between 23.1 and 26, or <18 kg given a BMI between 26.1 and 29, or <21 kg given a BMI >29 (19). We defined weight loss as a drop in body weight of more than 5 percent since the last examination at the research center. We coded participants as exhausted when they stated that the CES-D “I could not get going” and “I felt that everything I did was an effort”, described their feelings the last week “frequently” or “mostly”. Based on questionnaires with wide-ranging questions regarding physical activities and leisure time, we calculated the kilocalories per week as a sum score of the activities weighted by their intensity. When this sum value was <383 kcal per week in men and <270 in women, we defined them as having low physical activity. Slow walking speed was measured using a 5.79-m long walkway (GAITRite™ Platinum; CIR systems, Sparta, NJ: 4.88-m active area; 120-Hz sampling rate). We defined participants as slow walkers when their velocity was <0.76 m/s and their height was > 173 cm for men or > 159 cm for women, or otherwise when their velocity was <0.65 m/s. We did not assign a value for slow walking speed when the gait speed measurement was more than six months apart from the blood drawl of our participants.

We did not have information on all five domains in all participants. We only assigned an FP score to participants of whom we had information on at least three of the above-mentioned domains. We assigned an FP-score by the number of fulfilled frailty criteria divided by the number of assessed frailty criteria. Afterward, we multiplied the outcome by 5 to make the scores of participants with a different number of assessed frailty domains comparable. For the logistic regression analysis, we classified participants with a FP-score ≥2 as frail. We deviated from the in literature described FP ≥3 cut-off as our study population is very healthy.

#### Continuous frailty

Continuous frailty uses the same measurements as the frailty phenotype(20). However, instead of using discrete cut-offs and treating all domains equally, continuous frailty assigns different weights to the different domains and treats all measurements as continuous. The residual of a linear regression of sex and BMI on handgrip strength was used as the measure for low handgrip strength, whereas the residual of a linear regression of sex and height was used as the measure of gait speed. Weight loss was defined as the percentage of weight loss since the last visit to our research center. Exhaustion was measured as a sum score ranging from 1 to 12 based on the agreement of the participants with the statements “I could not get going” and “I felt that everything I did was an effort” during last week. When the participants answered “rarely/none of the time”, we coded this as 0.5, “some or a little of the time” was coded 1.5, “a moderate amount of time” was coded 3.5, or “most of the time” was coded 6. The total amount of kcal of physical activity was used for the domain of low physical activity.

To calculate continuous frailty, we subtracted the mean value per dataset from the observed value per participant and divided this by the standard deviation. Afterward, using the earlier weights developed in the Cardiovascular Health Study, we calculated the continuous frailty score(20).

#### Tilburg Frailty Indicator

The Tilburg Frailty Indicator (TFI) is a self-reported frailty questionnaire(22). We did not have the original questionnaire in our study population. Therefore, we selected the best possible proxy for each question. The proxies were based on questionnaires, observations during the interviews, and weight change was measured as a weight change of more than five percent since the last visit to the research center. A more detailed description of the proxies can be found in Appendix 4: Supplementary Table II. Unfortunately, we did not have a proper proxy for the question “Are you able to cope with problems well and the question “Do you receive enough support from other people?”. We included solely people who had at least information on ten of the thirteen other questions in the analyses. We divided the sum of the scores of all questions by the number of questions answered by a person and multiplied by thirteen to make the results comparable between participants. A score of four or higher was defined as being frail

#### Multidimensional Prognostic Index

The Multidimensional Prognostic Index (MPI) is a risk score for predicting 1-year mortality in older individuals based on the different domains covered by the Comprehensive Geriatric Assessment (CGA). Traditionally the MPI quantifies these domains based on the number of activities a person can undertake independently measured by the Katz’s Activities of Daily Living (ADL), as well as the Lawton’s Instrumental Activities of Daily Living (IADL), the Cumulative Illness Rating Scale Comorbidity Index (CIRS-CI), Exton Smith Scale (ESS), Nutritional Assessment Short Form (MNA-SF), the Short Portable Mental Status Questionnaire (SPMSQ), the medication intake and living situation of the participant(23).

We did not have the Katz’s Activities of Daily Living Scale, SPMSQ, nor the MNA-SF. Instead of using the ADL, we calculated the functional activities based on the Stanford Health Assessment Questionnaire (HAQ), we used the MMSE instead of the SPMSQ, and we replaced the MNA-SF with our own risk of undernutrition score. This undernutrition score consisted of the BMI categories as scored in the MNA-SF, quartiles of calorie intake and diet quality based on Food Frequency questionnaires, and a question if participants experienced difficulties eating. With the help of a geriatrician, we established the CIRS-CI and ESS based on proxies. Unfortunately, we did not have questions regarding incontinence. Therefore, the ESS was based on four instead of five questions. When participants missed more than two IADL questions, they did not get a score assigned for this domain. When one or two questions were missing, the sum of the answered questions was divided by the number of answered questions and then multiplied by eight to make all scores comparable. We used the same procedure in the case of our Nutri-score, the HAQ, and ESS.

Afterward, all domains were assigned a 0 indicating low risk of mortality or longer hospitalization, 0.5 indicating a moderate risk of mortality or more extended hospitalization, or 1 indicating high risk of mortality or hospitalization. We based our risk assignments on previous studies [31,37–40]. When people lacked information on more than two domains, we did not assign them an MPI score. Otherwise, we divided the sum of the different domains by the number of domains on which we had information. This resulted in an MPI risk score ranging between 0 and 1. Traditionally the MPI is categorized as low-risk, moderate-risk, and high risk of dying within a year. As we used the MPI in the general population, we decided to use the MPI continuously to avoid the loss of information. In the logistic regression, we used 0.33, the traditional cut-off for moderate risk as ‘frail’, as in our healthy population, we did not have participants with an MPI-score above 0.66.

### Online-Only Text 2

Supplementary Table I. Proxies used to calculate Tilburg Frailty Indicator.

| Question in Tilburg Frailty Indicator questionnaire | Rotterdam Study proxy |
| --- | --- |
| Do you feel physically healthy?  Yes (0)/No (1) | How do you rate your own health compared to other people you age? Better (0)/The same (0)/Worse (1)  EQ5D question 4: pain/complaints  I have no pain or discomfort (0)/ I have slight pain or discomfort (0)/ I have moderate pain or discomfort (0)/ I have severe pain or discomfort (1)  When at least one question is 1 🡪 (1). |
| Have you lost a lot of weight recently without wishing to do so? (‘a lot’ is: 6 kg or more during the last six months, or 3 kg or more during the last month)  Yes (1)/No (0) | Measured weight loss since last visit  >5% (1), <=5% (0) |
| Do you experience problems in your daily life due to difficulty in walking?  Yes (1)/No (0) | EQ5D question 1: mobility I have no problems in walking about.(0)/I have some problems in walking about (0)/ I am confined to bed (1) |
| Do you experience problems in your daily life due to difficulty maintaining your balance?  Yes (1)/No (0) | Stanford Health Assessment Questionnaire (15)  Are you able to stand up from an armless chair? Without any difficulties (0)/With some difficulties (0)/With a lot of difficulties (1)/Unable to do (1);  Are you able to get on and off the toilet? Without any difficulties (0)/With some difficulties (0)/With a lot of difficulties (1)/Unable to do (1);  When at least one question is 1 🡪 (1). |
| Do you experience problems in your daily life due to poor hearing?  Yes (1)/No (0) | Observations during interview filled in by interviewer.  Hearing of the participant: No signs of hearing loss (0)/hearing loss, needed to talk loudly (1)/Almost complete hearing loss, communication disturbed (1);  In participants without a hearing aid: Do you experience hearing loss? No, I never miss a beat (0)/Yes, I sometimes can’t follow what is said (1)/Yes, I often can’t follow what is said (1)/Yes, I almost never follow what is said (1);  In participants with a hearing aid: Do you experience hearing loss even when wearing you hearing aid? No, I never miss a beat (0)/Yes, I sometimes can’t follow what is said (1)/Yes, I often can’t follow what is said (1)/Yes, I almost never follow what is said (1);  Are you able to follow a conversation involving more than three persons? (Almost) never (1)/Sometimes (1)/Often (1)/(Almost) always (0);  Do you avoid events (such as birthdays) due to you hearing? (Almost) never (0)/Sometimes (1)/Often (1)/Almost always (1);  When one of these questions is 1 🡪 (1). |
| Do you experience problems in your daily life due to poor vision?  Yes (1)/No (0) | Observations during interview filled in by interviewer. Vision of the participant: No signs of vision loss (0)/Vision loss (1)/Nearly blind (1) |
| Do you experience problems in your daily life due to lack of strength in your hands?  Yes (1)/No (0) | Stanford Health Assessment Questionnaire (15)  Do you have problems with eating? Do you experience problems cutting meat or bread or lifting a full glass of milk to your mouth? Without any difficulties (0)/With some difficulties (cutting meat) (1)/With a lot of difficulties (lifting a full glass of milk) (1)/Unable to do (receives help eating) (1);  Are you able to open a new milk carton? Without any difficulties (0)/With some difficulties (0)/With a lot of difficulties (1)/Unable to do (1);  Are you able to turn faucets on and off? Without any difficulties (0)/With some difficulties (0)/With a lot of difficulties (1)/Unable to do (1);  Are you able to comb your hair and/or do your make-up? Without any difficulties (0)/With some difficulties (0)/With a lot of difficulties (1)/Unable to do (1);  Are you able to open car doors? Without any difficulties (0)/With some difficulties (0)/With a lot of difficulties (1)/Unable to do (1);  Are you able to open jars which have previously been opened? Without any difficulties (0)/With some difficulties (0)/With a lot of difficulties (1)/Unable to do (1);  Are you able to hold a pen and pencil? Without any difficulties (0)/With some difficulties (0)/With a lot of difficulties (1)/Unable to do (1);  When one of these questions is (1) 🡪 (1) |
| Do you experience problems in your daily life due to physical tiredness?  Yes (1)/No (0) | Pittsburgh Sleep Quality Index (PSQI)  During the past month, how much of a problem has it been for you to keep up enough energy to get things done? No problem at all (0)/Only a very slight problem (0)/Somewhat a problem (1)/A lot of problem (1). |
| Do you have problems with your memory?  Yes (1)/Sometimes (0)/No (0) | Observations during interview filled in by interviewer. Memory problems of the participant: No signs of memory problems (0)/Yes, moderate memory problems and/or not all the time (0)/Yes, severe memory problems or continue memory problems (1) |
| Have you felt down during the last month?  Yes (1)/Sometimes (1)/No (0) | CES-D (17)  In the last week, I felt that I could not shake off the blues even with the help of my family or friends. Rarely or none of the time [0-1 days] (0)/Some or a little of the time [1-2 days] (1)/Occasionally or a moderate amount of the time [3-4 days] (1)/Most or all of the time [5-7 days] (1);  In the last week, I felt depressed. Rarely or none of the time [0-1 days] (0)/Some or a little of the time [1-2 days] (1)/Occasionally or a moderate amount of the time [3-4 days] (1)/Most or all of the time [5-7 days] (1);  When one of these questions is (1) 🡪 (1) |
| Have you felt nervous or anxious during the last month?  Yes (1)/Sometimes (1)/No (0) | CES-D (17)  In the last week, I felt fearful. Rarely or none of the time [0-1 days] (0)/Some or a little of the time [1-2 days] (1)/Occasionally or a moderate amount of the time [3-4 days] (1)/Most or all of the time [5-7 days] (1). |
| Do you live alone?  Yes (1)/No (0) | How many people are part of your household? 1 (1) 1< (0) Who do you live with? I live alone (1)/My spouse/partner (0)/Children (0)/Brother(s)/sister(s) (0)/Other people namely … (0) When one of these questions is (1) 🡪 (1) |
| Do you sometimes miss having people around you?  Yes (1)/Sometimes (1)/No (0) | CES-D (17)  Last week, I felt lonely. Rarely or none of the time [0-1 days] (0)/Some or a little of the time [1-2 days] (1)/Occasionally or a moderate amount of the time [3-4 days] (1)/Most or all of the time [5-7 days] (1). |

Supplementary Table II. Proxies used to calculate Multidimensional Prognostic Index

| Question in Multidimensional Prognostic Index | Rotterdam Study proxy |
| --- | --- |
| Number of active functional activities in the Katz’s Activities of Daily Living (ADL)  6-5 (0); 4-3 (0.5); 2-0 (1). | Number of active functional activities based on the Stanford Health Assessment Questionnaire (HAQ)  8-6 (0); 5-4 (0.5); 3-0 (1). |
| Number of active functional activities in the Instrumental ADL (IADL)  8-6 (0); 5-4 (0.5); 3-0 (1). | Number of active functional activities in the Instrumental ADL (IADL)  8-6 (0); 5-4 (0.5); 3-0 (1). |
| Short Portable Mental Status Questionnaire (SPMSQ)  0-3 (0); 4-7 (0.5); 8-10 (1). | Mini-Mental State Examination (MMSE)  30-28 (0); 27-25 (0.5); 24> (0). |
| CIRS-CI  0 (0); 1-2 (0.5); 3≤ (1). | CIRS-CI  0 (0); 1-2 (0.5); 3≤ (1). |
| Mini Nutritional Assessment (MNA)  ≥24 (0); 17-23.5 (0.5); <17 (1) | Do you have problems with eating? Do you experience problems cutting meat or bread or lifting a full glass of milk to your mouth? Without any difficulties ~ 1/With some difficulties (cutting meat) ~ 2/With a lot of difficulties (lifting a full glass of milk) ~ 3/Unable to do (receives help eating) ~ 4;  Diet Quality Score  1^st^ quartile ~ 1; 2^nd^ quartile ~ 2; 3^rd^ quartile ~ 3; 4^th^ quartile ~ 4;  Kcal intake  1^st^ quartile ~ 1; 2^nd^ quartile ~ 2; 3^rd^ quartile ~ 3; 4^th^ quartile ~ 4;  Body mass index  <19 ~ 1; 19-21 ~ 2; 21-23 ~ 3; 23 ≤ ~ 4;  When the sum these four questions is  ≥12 (0); 11-8 (0.5); 8> (1) |
| Exton Smith Scale (ESS)  Physical condition Good ~ 4; Fair ~ 3; Poor ~ 2: Very bad ~ 1;    Mental condition Alert ~ 4; Apathetic ~ 3; Confused ~ 2; Stupor ~ 1;  Activity Ambulant ~ 4; Walk-help ~ 3; Chair-bound ~ 2; Stupor ~ 1.  Mobility Full ~ 4; Slightly limited ~ 3; Very limited ~ 2; Immobile ~ 1.  Incontinent Not ~ 4; Occasionally ~ 3; Usually urine ~ 2; Doubly ~ 1.  Total score 16-20 (0); 10-15 (0.5); 9> (1). | Exton Smith Scale (ESS)  Physical condition  ~ mean of following three questions: 1. How do you rate your own health compared to other people you age? Better – 4/The same – 3/Worse – 2; 2. Hand grip (kg) by sex 4^th^ quartile – 4; 3^rd^ quartile – 3; 2^nd^ quartile – 2; 1^st^ quartile – 1. 3. Gait speed (m/s) by sex 4^th^ quartile – 4; 3^rd^ quartile – 3; 2^nd^ quartile – 2; 1^st^ quartile – 1. Mental condition Observations during the interview: The participant did not seem confused, appeared to have a clear consciousness, did not perseverated, had no memory problems, and did not wander ~ 4; The participant appeared to have some to severe memory problems or some or not constant confusion or appeared to wander or perseverated sometimes or not severely ~ 3; The participant seemed continue or severely confused or did not appear to have a clear consciousness or severely or continuously perseverated ~ 2; The participant seemed continue or severely confused and did not appear to have a clear consciousness ~ 1. Activity  Based on ADL Able to travel by themselves ~ 4; Not able to travel by themselves or having a lot of difficulties standing up from an armless chair or having some difficulties getting out of bed by themselves ~ 3; Not able to stand up from an armless chair or a lot of difficulties with getting out of bed by themselves ~ 2; Not able to get out of bed by themselves ~ 1.  Mobility Do you use a walking aid or a wheel chair? None ~ 4; Cane or tripod cane ~ 3; A walking frame with or without wheels ~ 2; Wheel chair ~ 1;  Total score 13-16 (0); 8-12 (0.5); 7> (1). |
| Number of medications  0-3 (0); 4-6 (0.5); 7≤ (1). | Number of medications  0-3 (0); 4-6 (0.5); 7≤ (1). |
| Social support network  Living with family (0); Institutionalized (0.5); Living alone (1). | Social support network  Living with family (0); Institutionalized (0.5); Living alone (1). |

### Online-Only References

18.	Folstein MF, Folstein SE, Mchugh PR. *“Mini-Mental State” A Practical Method for Grading the Cognitive State of Patients for the Clinician**. Vol 12. Pergamon Press; 1975.

### Online-Only Figures

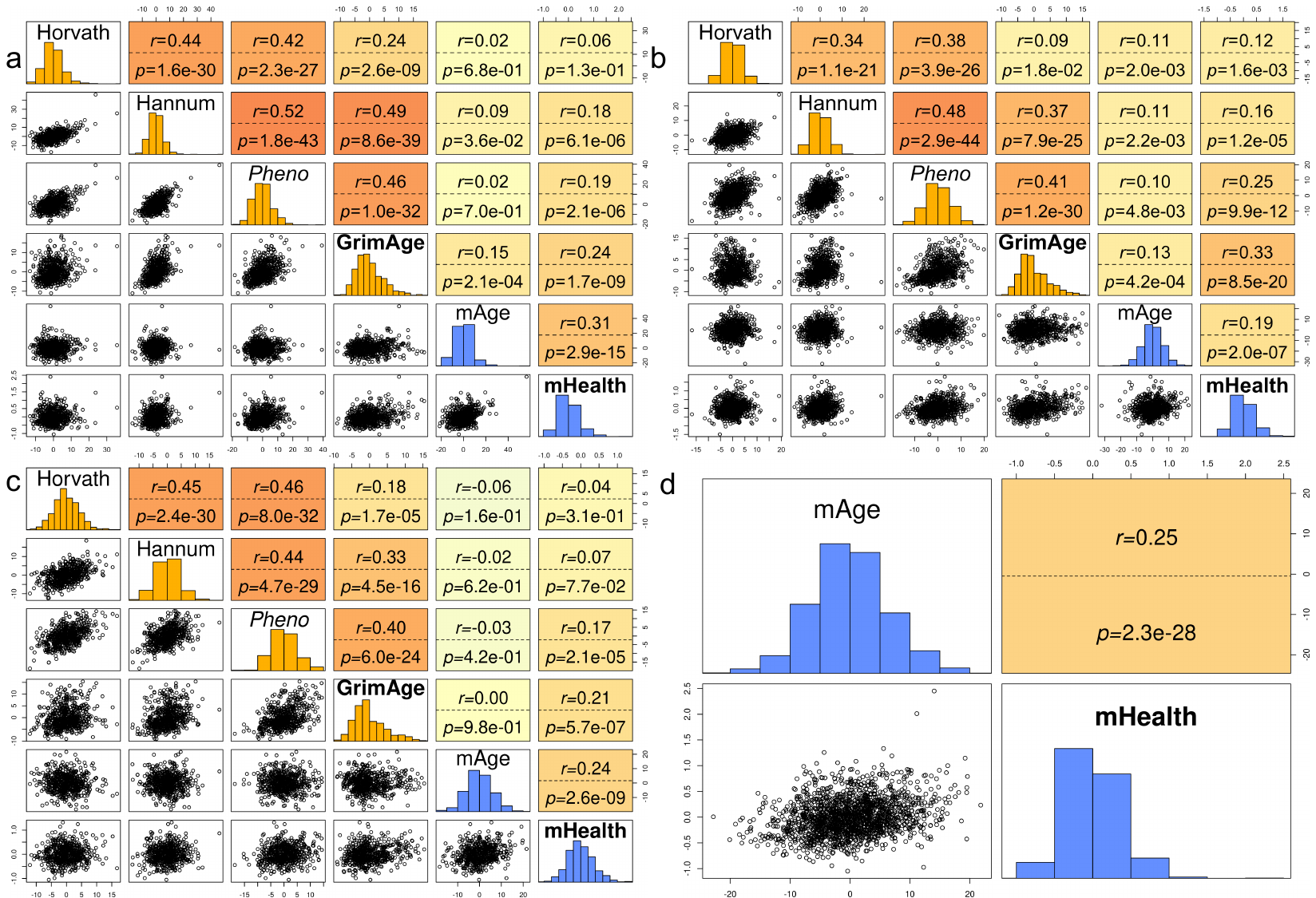

eFigure 1 Correlation of biological age biomarkers in Rotterdam Study subgroups and Leiden Longevity Study

Spearman’s correlation of the different biological aging biomarkers with epigenetic aging biomarkers in yellow, metabolomic-based aging biomarkers in blue. Values after r= represent Spearman’s rank coefficient; values after p= represent the p-value; the background color is darker for higher correlations. In: a. The Rotterdam Study 450K-subcohort (n=611); b. The Rotterdam Study EPIC-subcohort (n=736); c. The subcohort of the Leiden Longevity with information on all six epigenetic and metabolomic aging biomarkers (n=580); d. The entire Leiden Longevity Study study population (n=1,849).

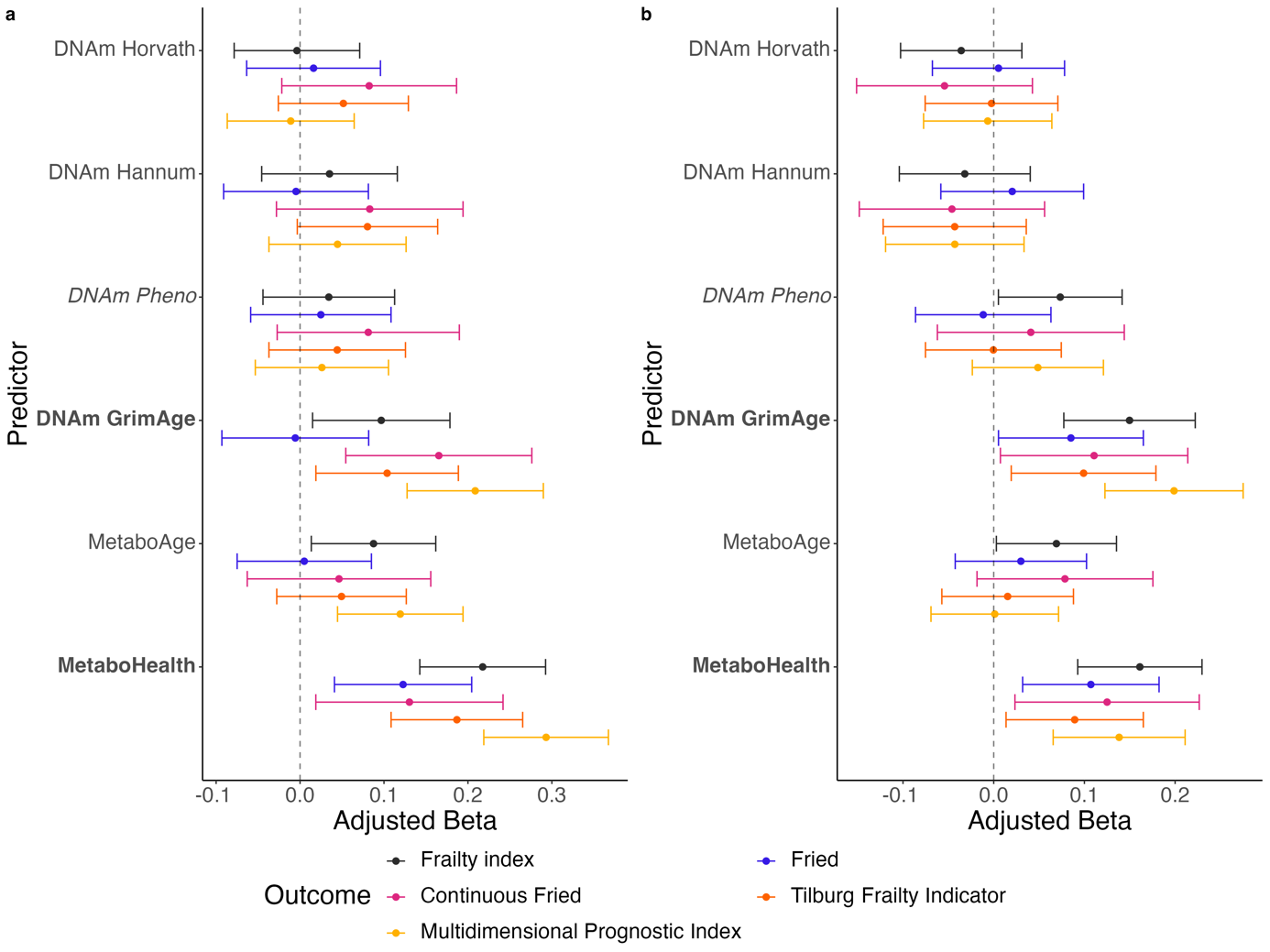

eFigure 2 Associations between biological aging biomarkers and frailty measures in Rotterdam Study subcohorts

**a.** Associations of standardized biological aging biomarkers with standardized FI(*n=*608), FP(*n=*606), Cont.(*n=*350), TFI(*n=*608), and MPI(*n=*609) based on linear regression analyses in all participants of whom data on biological aging biomarkers and frailty scores were available in Rotterdam Study’s 450K-subcohort. The figure represents the adjusted betas and 95%-confidence intervals. **b.** Associations of standardized biological aging biomarkers with standardized frailty index(*n=*733), Fried(*n=*733), Cont.(*n=*398), TFI(*n=*731), and MPI(*n=*735) based on linear regression analyses in all participants of whom data on biological aging biomarkers and frailty scores were available in Rotterdam Study’s EPIC-subcohort. The figure represents the adjusted betas and 95%-confidence intervals.

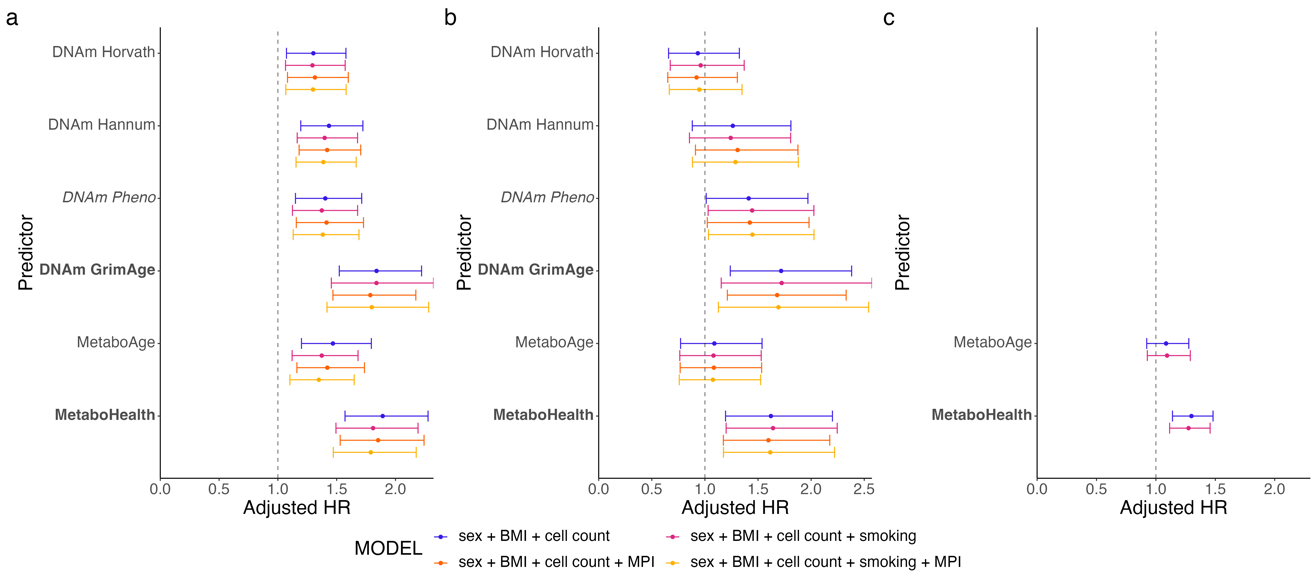

eFigure 3 Aging predictors and their univariable risk of all-cause mortality per standard deviation in the Rotterdam Study subcohorts and Leiden Longevity Study overall population

Risk of all-cause mortality per standard deviation increase of the aging predictors in **a.** the 450K-subgroup (n=609); **b.** the EPIC-subgroup (n=727); and c. the overall study population of the Leiden Longevity Study (n=1,849). BMI indicates body mass index; HR, hazard ratio; and MPI, multidimensional prognostic index.
